# Global lymphatic filariasis post-validation surveillance activities in 2024: A systematic review protocol

**DOI:** 10.1101/2024.12.14.24319037

**Authors:** Holly Jian, Harriet Lawford, Angus McLure, Colleen Lau, Adam Craig

## Abstract

**Introduction:** Lymphatic filariasis (LF) is a neglected tropical disease caused by infection with parasitic worms, spread by mosquitoes. In countries where LF is validated as eliminated as a public health problem by the World Health Organization (WHO), post-validation surveillance (PVS) is required to ensure recrudescence has not occurred and verify the sustained elimination of transmission. However, it is unclear what PVS strategies should be applied, how PVS strategies should be tailored to meet country capacity and need, and whether currently used approaches align with upcoming WHO guidelines.

**Objectives:** This study will aim to review available evidence on PVS implementation in countries previously endemic for LF; examine barriers and facilitators to PVS implementation; critique alignment in PVS activities with international guidelines; and identify knowledge gaps in PVS implementation that may be addressed through further research.

**Methods:** We will search four databases (PubMed, Scopus, Embase and Web of Science) for peer-reviewed literature and the WHO Institutional Repository for Information Sharing (IRIS) database for grey literature. Documents published between January 1, 2007 and November 5, 2024 will be included. Two reviewers will independently screen studies based on a priori inclusion and exclusion criteria. The quality of included studies will be assessed using the Joanna Briggs Institute Critical Appraisal Checklist, and deductive content analysis will be conducted to synthesise data. The study will also examine alignment with upcoming WHO PVS guidelines.

**Conclusion:** This review will systematically collate and analyse available literature on PVS of LF, which, to our knowledge, has not yet been conducted. Our study will synthesise knowledge in this field and provide an evidence base which may be used to guide the design of future PVS strategies.

This protocol has been registered in PROSPERO (registration ID: <u>CRD42024618436</u>).

## Introduction

Lymphatic filariasis (LF) is a neglected tropical disease caused by infection from three species of thread-like parasitic worms: *Wuchereria bancrofti, Brugia malayi* and *B. timori. W. bancrofti* causes 90% of infections worldwide and is spread primarily by *Aedes, Anopheles* and *Culex* genus mosquitoes. The typical vector for *Brugia* spp. filariasis are mosquito species in the genera *Mansonia* and *Aedes* [1, 2].

Lymphatic filariasis is endemic in 72 countries worldwide [3]. Chronic infection can lead to lymphoedema (tissue swelling) or elephantiasis (skin and tissue thickening) of limbs and hydrocoele (scrotal swelling) [4]. The resulting disfigurement and disability can lead to physical impairment and social stigmatisation, and individuals are often ostracised and lose employment [5]. In 1998, prior to the establishment of the Global Programme for the Elimination of Lymphatic Filariasis (GPELF), chronic LF had an estimated disability-adjusted life-years (DALY) burden of 5.25 million and an estimated economic burden of USD 2.5 billion annually, largely due to disability and loss of employment. In 2019, the burden was reported to have more than halved with burden being estimated at 1.63 million DALYs [6, 7].

The GPELF was launched in 1998 and is one of the largest global public health interventions ever mounted. The programme aims to eliminate LF as a public health problem through multiple rounds of mass drug administration (MDA), as well as managing morbidity and preventing disability among those already infected [8, 9]. Regional and national programmes were later established, recognising that diverse epidemiological profiles and contextual factors required tailored strategies for successful implementation. These include the establishment of the Pacific Programme for the Elimination of Lymphatic Filariasis (PacELF) in 1999 [1], the South-East Asia regional programme to eliminate lymphatic filariasis in 2000 [10], and various national programmes across the World Health Organization (WHO) Regional Office for Africa [11].

The elimination of LF as a public health problem is defined by WHO as reducing prevalence of infection to below defined target thresholds [12]. Once targets have been reached, countries are validated by WHO as having eliminated LF as a public health problem. The validation process requires the preparation and submission of a dossier proving the sustained reduction of LF prevalence nationally. This is based on initial surveys in high-risk sentinel sites that show the prevalence of infection is below the target threshold in populations aged over five years of age, known as pre-transmission assessment surveys (pre-TAS). These are then followed by a transmission assessment survey (TAS1) showing that the prevalence of infection is below the target threshold in a sample of children under seven years. After MDA stops, subsequent surveys must show sustained low prevalence of infection in children aged under seven for two years (TAS2) and four years (TAS3) [13]. The thresholds used in these surveys are defined as an antigen or antibody prevalence level at which further transmission is unlikely, even in the absence of further rounds of MDA or other interventions. Thresholds vary depending on the predominant filarial species and vector in each country [14].

Following the validation of elimination of LF as a public health problem, WHO guidelines require that post-validation surveillance (PVS) be conducted [13]. PVS is ongoing surveillance following elimination, with the aim of ensuring recrudescence has not occurred. A secondary aim of PVS is to detect and provide care for persons affected by lymphoedema and hydrocoele [15].

Provisional guidelines suggest that PVS should be continued for at least ten years after validation to monitor for recrudescence. Evidence emerging from the Pacific islands and Sri Lanka indicates that, even after the criteria for elimination as a public health problem have been met, local LF transmission can continue. Where appropriate PVS is not implemented, continued transmission may result in re-establishment of transmission and threatening the successes of many years of MDA [16–18].

The WHO *Road Map for Neglected Tropical Diseases 2021–2030* aim that all 72 LF-endemic countries will no longer require MDA and will be implementing PVS or post-MDA surveillance by 2030. Additionally, the Road Map aims for at least 80% of LF endemic countries (58/72) to have been validated for the elimination of LF as a public health problem by 2030 [19].

China, in 2007, became the first country to be validated by WHO as having eliminated LF as a public health problem. Since then (up to November 2024), 21 other countries have met the epidemiological threshold, prepared and submitted a dossier and been validated by WHO as having eliminated LF. Twelve others have reduced prevalence levels to below the epidemiological threshold for elimination but are still completing TAS. The disease remains endemic in 39 countries that still require MDA [3, 8, 20].

**Figure 1:**
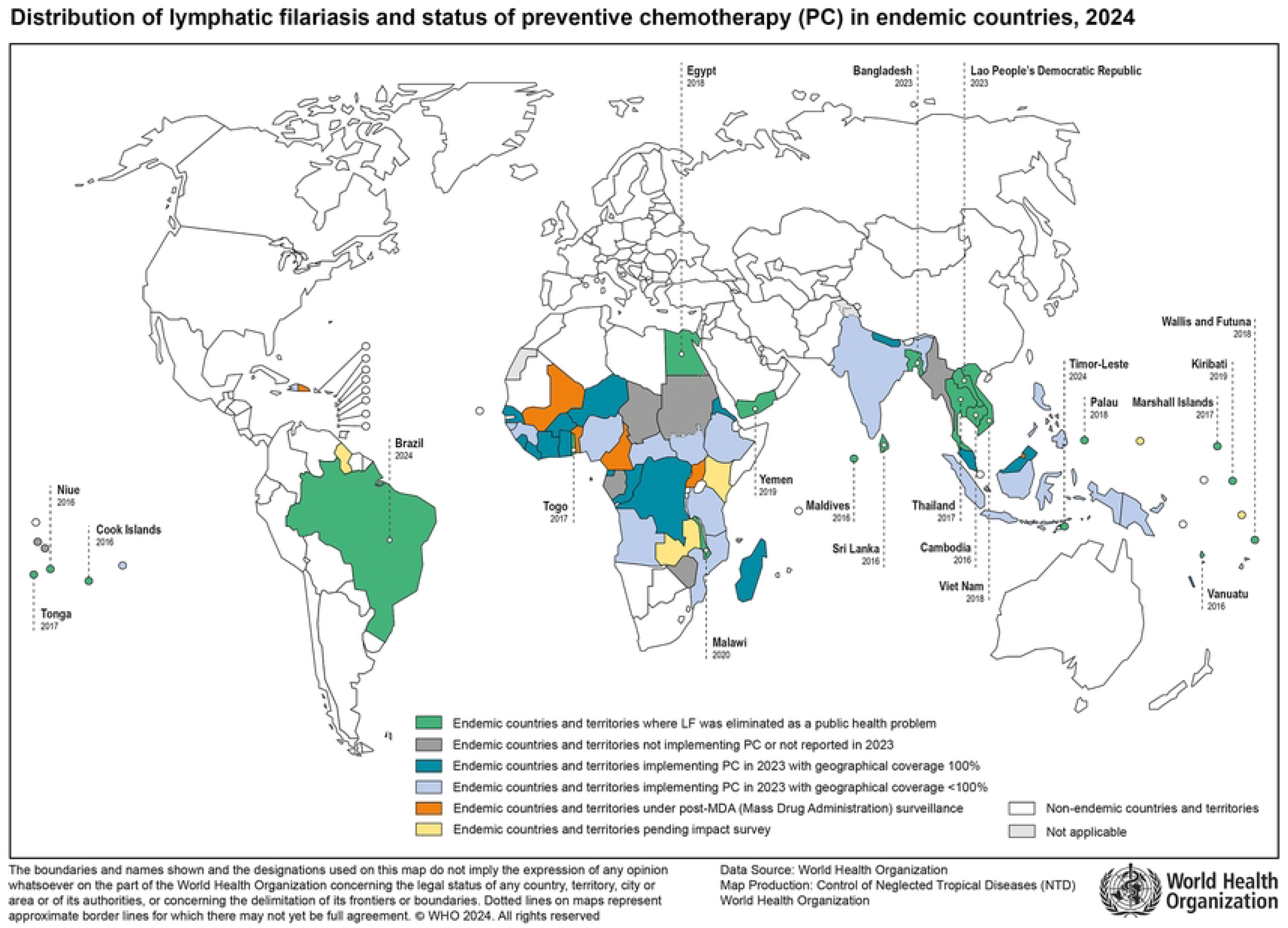
Distribution of lymphatic filariasis and status of preventative chemotherapy (PC) in endemic countries, 2024. At the time of writing (November 2024), WHO guidelines for PVS had been drafted and presented at international meetings but had not officially been released. Based on the content presented, we expect that the guidelines will recommend countries implement two of the following four PVS strategies: health-facility based screening; integration of PVS into existing population surveys; targeted LF surveillance of high-risk groups; and molecular xenomonitoring [16].

The implementation of effective PVS requires a robust understanding of countries’ unique geographic and epidemiological profile, operational context, and challenges and enablers to PVS implementation. Importantly, experiences in one country may inform, but should not dictate, approaches taken by another; context, capacity, and need will inevitably differ. Given this, the aims of this systematic review are to identify and synthesise evidence on PVS activities globally, establishing an evidence base on which future guidelines and strategies may draw. Specifically, the objectives of this review are to:

1. Profile post-validation or post-elimination surveillance activities, or lack thereof, in countries previously endemic for LF that have been validated by the WHO as having eliminated LF
2. Examine barriers and facilitators to the implementation of PVS strategies, and how these vary by context.
3. Identify alignment of PVS activities with upcoming WHO guidelines
4. Identify knowledge gaps in PVS implementation methods that may be addressed through further operational research.

## Methods

This study will be conducted in line with the PRISMA guidelines.

### Research question

The Population, Intervention/Exposure, Comparator, Outcome and Time (PICOT) framework was used to develop research questions. As this review aims to scope and critique the PVS strategies being employed across several countries, we determined that identifying a Comparator was not required.

**Table 1:** PICOT components.

| PICOT |  |
| --- | --- |
| Population | Countries that have been validated by WHO as having eliminated LF:<br>Bangladesh, Cambodia, Cook Islands, Egypt, Kiribati, Lao People's Demographic Republic, Malawi, Maldives, Marshall Islands, Niue, Palau, Sri Lanka, Thailand, Togo, Tonga, Vanuatu, Vietnam, Yemen, Wallis & Futuna, Brazil and Timor-Leste |
| Exposure or intervention | Lymphatic filariasis |
| Outcome | Post-validation or post-elimination surveillance |
| Comparator | N/A |
| Time | 2007–2024 (inclusive) |

### Identifying studies

#### Information sources and search strategy

PubMed, Scopus, Embase and Web of Science will be used to search for peer-reviewed journal articles; the WHO Institutional Repository for Information Sharing (IRIS) database was used to search for grey literature reports published by WHO.

A search strategy using MeSH search terms, free text key words, and Boolean operators has been developed. Table 2 outlines the search strategy that will be used. Appendix A presents the search terms used across each literature database.

**Table 2:**
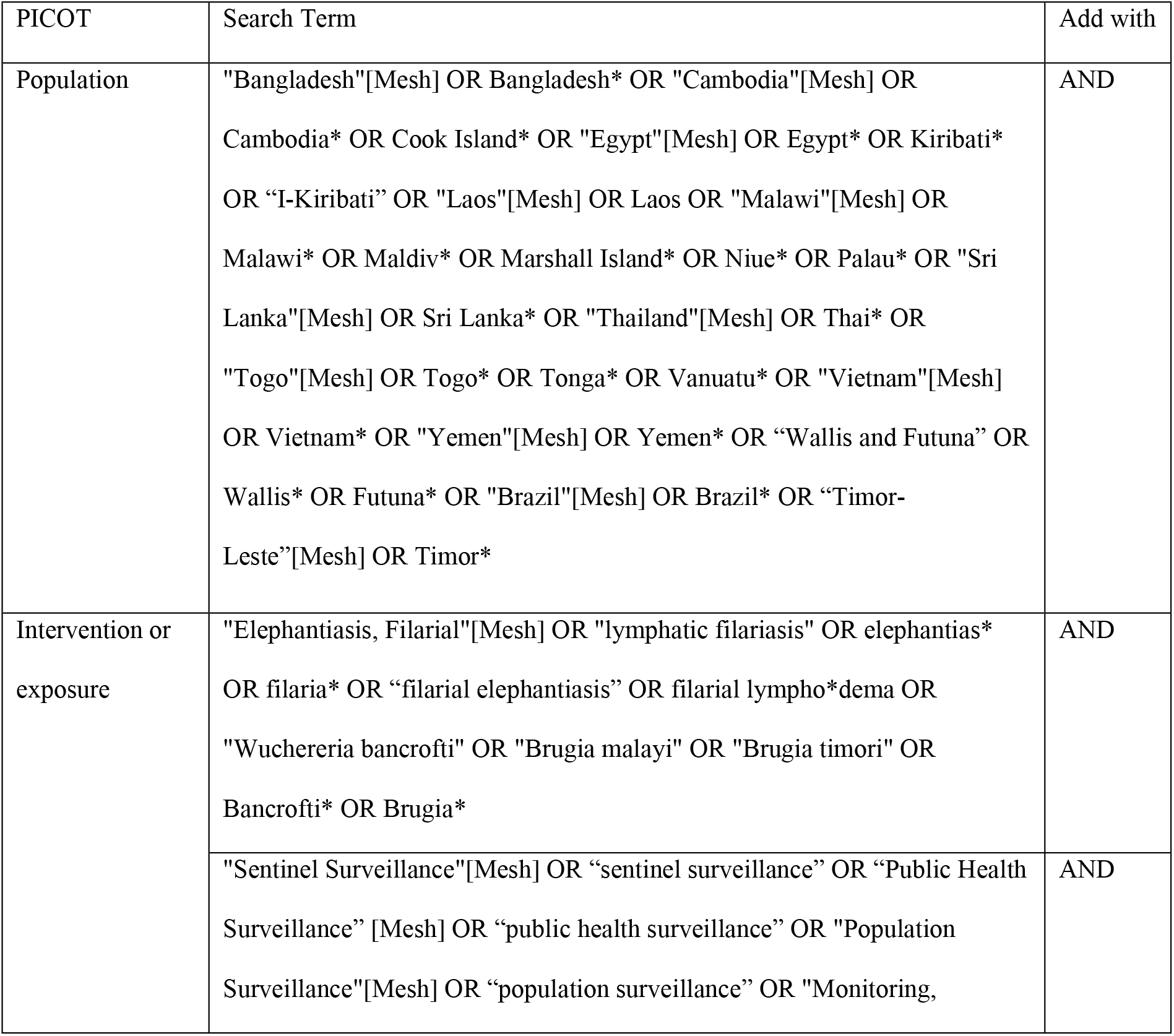
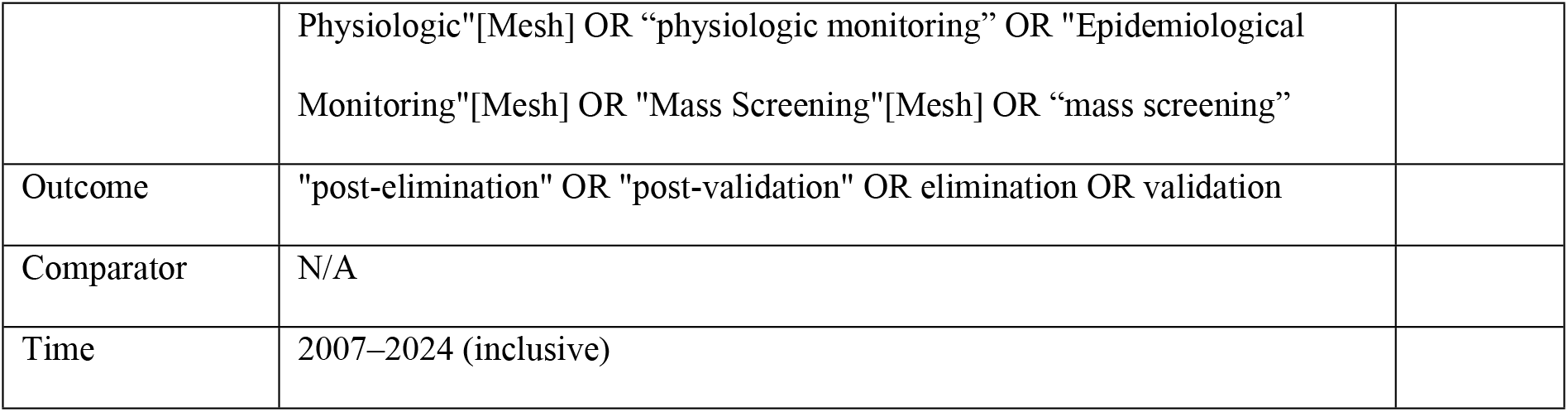
Search terms by PICOT.

| PICOT | Search Term | Add with |
| --- | --- | --- |
| Population | "Bangladesh"[Mesh] OR Bangladesh* OR "Cambodia"[Mesh] OR<br>Cambodia* OR Cook Island* OR "Egypt"[Mesh] OR Egypt* OR Kiribati*<br>OR "I-Kiribati" OR "Laos"[Mesh] OR Laos OR "Malawi"[Mesh] OR<br>Malawi* OR Maldiv* OR Marshall Island* OR Niue* OR Palau* OR "Sri<br>Lanka"[Mesh] OR Sri Lanka* OR "Thailand"[Mesh] OR Thai* OR<br>"Togo"[Mesh] OR Togo* OR Tonga* OR Vanuatu* OR "Vietnam"[Mesh]<br>OR Vietnam* OR "Yemen"[Mesh] OR Yemen* OR "Wallis and Futuna" OR<br>Wallis* OR Futuna* OR "Brazil"[Mesh] OR Brazil* OR "Timor-Leste"[Mesh] OR Timor* | AND |
| Intervention or<br>exposure | "Elephantiasis, Filarial"[Mesh] OR "lymphatic filariasis" OR elephantias*<br>OR filaria* OR "filarial elephantiasis" OR filarial lympho*dema OR<br>"Wuchereria bancrofti" OR "Brugia malayi" OR "Brugia timori" OR<br>Bancrofti* OR Brugia* | AND |
|  | "Sentinel Surveillance"[Mesh] OR "sentinel surveillance" OR "Public Health<br>Surveillance" [Mesh] OR "public health surveillance" OR "Population<br>Surveillance"[Mesh] OR "population surveillance" OR "Monitoring, | AND |
|  | Physiologic"[Mesh] OR "physiologic monitoring" OR "Epidemiological Monitoring"[Mesh] OR "Mass Screening"[Mesh] OR "mass screening" |  |
| Outcome | "post-elimination" OR "post-validation" OR elimination OR validation |  |
| Comparator | N/A |  |
| Time | 2007–2024 (inclusive) |  |

#### Data management, study screening and selection

Data management, study screening and selection will be performed with the aid of Covidence software (https://www.covidence.org/).

Search results from the selected databases will be imported into Covidence, and duplicates removed. Papers will be included if they:

- Describe PVS activities conducted between 2007–24 (inclusive) in a country or territory that has been validated by WHO as having eliminated LF.
- Were original research, activity reports, protocols or WHO grey literature describing population-level surveillance
- Were written in English, Portuguese, French or Chinese.
- Were published between January 2007 and November 2024.

Papers will be excluded if they:

- Describe only surveillance activities that occurred pre-validation by WHO of elimination of LF.
- Do not describe studies in a LF-eliminated country or territory.
- Do not describe surveillance activities (including diagnostic validation and modelling studies, letters, editorials and commentary articles).

Two researchers (HJ and HL) will independently screen the selected records using the above criteria by title and abstract. Where mismatches in screening assessments occur, HJ and HL will discuss and aim to find consensus; if required, a third reviewer (AC) will be consulted. If resolution cannot be found, a conservative approach will be taken and the article included in the review. Following title and abstract screening, HJ will conduct full-text screening.

Screening decisions will be documented using a PRISMA flow diagram.

### Data extraction

Data will be extracted by one researcher (HJ) using a Covidence data extraction form developed by the research team. Table 3 outlines the fields for which data will be extracted.

**Table 3:** Information to be extracted.

| Category | Item |
| --- | --- |
| Publication information | • Year of publication<br>• Country<br>• Funding sources |
| Epidemiology of LF | • Type of LF (*Wuchereria bancrofti*, *Brugia malayi*, *Brugia timori*)<br>• Species of LF vector (*Culex*, *Aedes*, *Mansonia*)<br>• Co-endemic diseases<br>• Year in which LF was validated as eliminated<br>• Prevalence of LF during<br>1. Endemic phase<br>2. Post-MDA surveillance<br>3. Post-validation surveillance<br>• High-risk or priority populations during elimination (e.g. historic hot spots) |
| Post-validation surveillance activities | • Years in which PVS activities were conducted<br>• Type of surveillance activities conducted<br>• Alignment with WHO guidelines (activity type, frequency)<br>• Sampling design (targeted, clustered, population-representative)<br>• Surveillance type (active, passive, sentinel)<br>• Type of testing (human antigen, antibody, and microfilaria; molecular xenomonitoring of mosquitoes)<br>• Other activities (including risk reduction and vector management) |
| Operational challenges or opportunities | • Funding sources<br>• Staffing<br>• Resources<br>• Technical capacity<br>• Social, political or economic drivers |

### Analysis

A narrative synthesis of PVS activities will be conducted. This will aim to describe PVS activities by country, their alignment with WHO guidelines, and specific challenges or enablers to their implementation. The review will examine differences or similarities between countries and aim to identify learnings that can be translated across settings.

A deductive content analysis approach will be employed, using the methodology outlined by Elo and Kyngas (2008). A deductive approach was selected due to its applicability in testing existing models in different contexts; in this case, this approach will be used to examine the application of PVS strategies recommended by WHO across countries endemic for LF. Deductive coding will be conducted using a constrained categorisation matrix based on Table 3, with a focus on identifying emerging themes relating to alignment with WHO guidelines, operational challenges and opportunities [21].

### Ethical considerations and dissemination of results

This review does not require ethical approval as it will use published data only. This study will be internally funded by The University of Queensland.

We anticipate publishing our results in a peer-reviewed publication. Additionally, we expect that the work will be presented at relevant international conferences and global and national LF elimination-related meetings.

### Quality assessment

Two reviewers will independently assess the quality of selected studies based on the eight criteria as described in the Joanna Briggs Institute critical appraisal checklist for use in reviews of cross-sectional studies (Appendix C). Each criterion will be given a score of 1 if a criterion is met or 0 if a criterion is not met, with a maximum possible score of 8. Studies scoring from 0 to 4 will be considered low quality, from 5 to 7 moderate quality, and from 8 to 9 high quality [22].

## Discussion

Our systematic review will synthesise the literature on PVS activities in countries that have been validated as having eliminated LF as a public health problem. It is anticipated that this review will bring together evidence on challenges and enablers in the implementation of these activities, as well as identify gaps that may require further operational research. Additionally, we anticipate that the review will assess whether what has been conducted in practice aligns with WHO PVS guidelines. The results of this review will help establish an evidence base to inform future PVS policies and programs.

The GPELF and its regional sub-programs have been recognised as successful public health interventions for the elimination of LF in 21 countries and the prevention of more than 3 million DALYs. However, as more countries achieve elimination of LF as a public health problem, there is a pressing need to ensure that any resurgence is detected in a timely way through the implementation of appropriate PVS. Without PVS, there remains the potential for LF recrudescence to go undetected, leading to further costly and preventable illness.

Each LF-endemic country has a unique epidemiological profile, opportunities, and constraints for the implementation of PVS which must be considered when designing national PVS strategies. As such, gathering evidence to inform on the selection and tailoring of PVS strategies is key for the ongoing success of LF elimination efforts. Understanding which PVS strategies have been employed and what the experience of implementation has been will provide valuable insights that others may draw on when designing their own PVS programs. Further, generating evidence as to whether what has been effective in practice aligns with existing guidelines will help tailor future recommendations.

Due to the diverse contexts in which PVS takes place, we expect there will be significant heterogeneity in the included studies. Additionally, without formal publication requirements, it is possible that countries conducting PVS may not publish data describing some or all of their activities, though we anticipate that some of these findings will be captured in the grey literature. Further, we acknowledge that valuable experiences and lessons learned will not have been captured in the scientific or grey literature and hence missed by this review. Recognition of this limitation opens opportunities for future research.

This review will map and evaluate the quality of the body of evidence on PVS of LF, which, to our knowledge, has not yet been conducted. Through collating and analysing literature on what strategies have been employed by previously endemic countries, this review will provide evidence on the operational feasibility of various PVS strategies, including those that will be recommended by WHO. Additionally, the review will identify knowledge gaps that require further research. We expect that our findings will assist and guide the design of future PVS strategies.

## Data Availability

No datasets were generated or analysed during the current study. All relevant data from this study will be made available upon study completion.

